# Prevalence and Molecular Characterization of Extended-Spectrum β-Lactamase-Producing *Escherichia coli* and *Klebsiella pneumoniae* in Clinical Enterobacterales from Dédougou, Burkina Faso

**DOI:** 10.1101/2025.06.25.25330325

**Authors:** Hiliassa Coulibaly, Dissinviel Stéphane Kpoda, Hervé Kafando, Oumar Traoré, Alix Bénédicte Kagambega, Alassane Halawen Mohamed, Robin Fréderic, Racha Beyrouthy, Dramane Ouédraogo, Abraham Ajayi, Dominique Bayala, Nicolas Barro, Richard Bonnet, René Dembélé

**Affiliations:** Laboratory of Molecular Biology, Epidemiology and Surveillance of Bacteria and Viruses Transmitted by Food, Ouagadougou BP 7021, Burkina Faso; Biomedical Laboratory of the Regional Hospital Centre of Dédougou, Dédougou BP 176, Burkina Faso; Microbiology and Microbial Biotechnology Laboratory (LAMBM), Ouagadougou BP 7021, Burkina Faso; Microbiology Laboratory of the General Reference Hospital (GRH), Niamey BP 12674, Niger; University Hospital Centre of Clermont-Ferrand, National Reference Centre (CNR) for Antibiotic Resistance, Clermont-Ferrand BP 63 000, France; Molecular Biology and Biotechnology Department, Nigerian Institute of Medical Research, Yaba, Lagos, Nigeria

**Keywords:** Prevalence, Antibiotic resistance, ESBL Enterobacterales, Molecular characterization, Dédougou

## Abstract

This cross-sectional study aimed to assess the frequency of Enterobacterales producing extended-spectrum ß-lactamases (ESBL) and to conduct a molecular characterisation of the ESBL resistance genes circulating in this region of Burkina Faso. Enterobacterales were isolated from clinical samples collected from the two main healthcare centres in Dédougou (Regional Hospital Center and Notre Dame de Bonne Délivrance Clinic). Isolates were identified using API 20E and confirmed by MALDI-TOF using a Vitek^®^ MS system. Antibiotic susceptibility testing was performed using the disk diffusion method. The ESBL genes and *E. coli* phylogenetic groups were characterized by polymerase chain reaction.

Among the 274 recovered *Enterobacterales* isolates, 148 (54%) were ESBL producers, predominantly *E. coli* (n = 112/194; 57.73 %) and *K. pneumoniae* (n = 24/54; 44.44 %). The *blaCTX-M-1* group genes were the most prevalent (88.97%; 121/136) among the ESBL-producing *Enterobacterales*, followed by the *blaCTX-M-9* group genes (11.03%; 15/136). All 112 ESBL-producing *E. coli* isolates were classified into four phylogenetic groups: A (n = 48/112, 42.86%), B1 (n = 9/112, 8.03%), B2 (n = 53/112, 47.32%), and D (n = 2/112, 1.79%).

The results of this study highlight a concerning spread of ESBL genes in this region of Burkina Faso. Public health efforts should prioritize education both for the general public and healthcare professionals, along with surveillance. Promotion of appropriate and restricted use of antibiotics is advocated to limit the spread of multi-resistant bacteria at the local level.

## Introduction

β-lactam antibiotics are the primary treatment for infections caused by *Enterobacterales*. However, in recent years, increasing resistance to these antimicrobials has emerged, posing significant public health concern. The primary resistance mechanism involves the production of β-lactamase enzymes, which hydrolyze the β-lactam ring, rendering the antibiotics ineffective Extended-spectrum β-lactamases (ESBLs) are among the most concerning resistance enzymes, characterized by extreme diversity due to their continuous evolution [1]. The observed multidrug resistance may result from the improper use and inadequate monitoring of β-lactam antibiotics, as well as poor hygiene in both community and hospital settings [2]. Today, the spread of ESBL-producing *Enterobacterales* has driven the epidemiological dissemination of *CTX-M*-type resistance genes, with *Escherichia coli* (*E. coli*) and *Klebsiella pneumoniae* (*K. pneumoniae*) being the primary species involved [3]. These resistance genes are typically plasmid-borne and often harbored alongside other antibiotic resistance genes, such as those conferring resistance to aminoglycosides and fluoroquinolones [4]. This leads to prolonged hospital stays, socio-economic challenges, and increased morbidity and mortality in affected patients [5]. This issue, recognized as a major global public health concern, has been reported in European countries [4,6], African countries [3,7,8], and particularly in Burkina Faso, especially in major cities such as Ouagadougou and Bobo-Dioulasso [9–11]. However, despite being one of the country’s largest regions known as the “attic of Burkina Faso” due to its agricultural productivity the Boucle du Mouhoun region, with its high level of urbanization, has not yet been the focus of such studies. Therefore, this study aimed to investigate the molecular epidemiology of ESBL-producing *E. coli* and *K. pneumoniae* isolates collected from clinical samples from 2 hospitals in Boucle of Mouhoun.

## Methods

### Ethics approval and consent to participate

The study was conducted according to the guidelines of the Declaration of Helsinki of 1975, as revised in 2013 and approved by The Ethical Committees of the hospital authorities of the “Régional Hospital Centre of Dédougou” (Reference Code: N°2022-124/MS/SG/CHR-DDG/DG/DSIO, 21 April 2022) and by The National Ethical Committees of Burkina Faso (N° 2022-05-097, 4 May 2022). Informed consent verbal was obtained from the adult patients themselves or accompanying adults of study participants prior to any collection of epidemiological data and samples. This study was reported in line with the STROBE (Strengthening the Reporting of Observational Studies in Epidemiology) guidelines for cross-sectional studies [34].

### Sites, study period and sampling

This cross-sectional study was conducted from 15 May 2022 to 31 December 2023. Routine sample collection was carried out in the two most frequented healthcare facilities in Dédougou (DDG): the Regional Hospital Centre (RHC) and the Notre Dame of Bonne Délivrance Clinic (NDBDC). It began on 15 May 2022 and ended on 30 April 2023. The target population consisted of patients attending these centres for Cyto-Bacteriological Examinations (CBE), regardless of sex or age. Any patient with a well-informed report and samples under the conditions indicated for an CBE was considered, and any patient whose samples were not under these conditions or who brought a sample for a study other than the cytobacteriological study was excluded from the study. A total of 1,845 clinical samples, including urine (n = 963), pus (n = 298), blood (n = 260), vaginal swabs (n = 151), urethral swabs (n = 20), and semen (n = 153), were consecutively collected from these facilities. The collected samples were properly labeled and immediately transported to the bacteriology section for analysis.

### Microbial culturing and identification

Gram-negative Bacilli (GNB) were isolated from clinical samples following the laboratory’s sample analysis protocol. After 18 to 24 hours of incubation at 37°C, *Enterobacterales* colonies were suspected based on an oxidase test. The suspected colonies were purified by inoculating them onto Müller-Hinton (MH) agar (Laboratorios Conda S.A) and incubating for 18 to 24 hours at 37°C. Bacterial identification was performed using the API 20E gallery (BioMérieux, Marcy-l’Étoile, France). After identification, isolates were plated onto MH agar, incubated for 18 to 24 hours at 37°C, then harvested and stored in CliniswabTS solid transport media (Italy) at room temperature before being transferred to the National Reference Center (NRC) in Clermont-Ferrand, France. *E. coli* and *K. pneumoniae* isolates were subcultured onto CPSE chromogenic agar (CHROMID-CPS-Elite, BioMérieux, France), and identification was confirmed by MALDI-TOF (Matrix-Assisted Laser Desorption/Ionization Time-of-Flight) using the VITEK® MS system (BioMérieux, France).

### Antibiotic susceptibility testing

Antibiotic susceptibility testing of bacterial isolates was performed on Müller-Hinton agar (Bio-Rad, France) using the disk diffusion method, following the recommendations of the Antibiogram Committee of the French Microbiology Society/European Committee on Antimicrobial Susceptibility Testing (CA-SFM/EUCAST, 2019). The 120 mm Müller-Hinton agar plates (Bio-Rad, France) were inoculated by swabbing with a 0.5 McFarland bacterial suspension, measured using a densitometer (DENSIMAT, BioMérieux, France), before dispensing fourteen antibiotic discs. After 18 to 24 hours of incubation at 37°C, the ADAGIO^TM^ system (Bio-Rad, France) was used to scan the diameters of the inhibition zones. The tested antibiotics included ampicillin (10 µg), amoxicillin + clavulanic acid (20/10 µg), cefoxitin (30 µg), cefotaxime (5 µg), ceftazidime (10 µg), aztreonam (30 µg), imipenem (10 µg), ertapenem (10 µg), gentamicin (10 µg), nitrofurantoin (100 µg), ciprofloxacin (5 µg), fosfomycin (200 µg), and trimethoprim + sulfamethoxazole (1.25/23.75 µg). Extended-spectrum β-lactamase (ESBL) production was assessed using the double-disc synergy test with amoxicillin + clavulanic acid and a third-generation cephalosporin [12]. After overnight incubation at 37°C, the presence of a champagne cork or keyhole pattern was considered indicative of ESBL production [13]. *E. coli* ATCC 25922 was used as a quality control strain to verify the reliability of the antibiotic discs before use.

### Phylogenetic Characterization and molecular profiling of multidrug-resistant *E. coli* isolates with ESBL gene identification

The genomic DNA was extracted from ESBL-producing *E. coli* and *K. pneumoniae* isolates using the boiling method, as previously described [14]. The concentration and purity of the extracted DNA were determined using a Nanodrop spectrophotometer (Implen P360 Nanophotometer) and stored at –20°C. In total, 112 ESBL-producing *E. coli* and 24 ESBL-producing *K. pneumoniae* isolates, exhibiting very similar resistance phenotypes, had their DNA extracted for molecular analysis.

The different phylogenetic groups of *E. coli* isolates were identified using the PCR-based method described by Clermont et *al*. [15,16]. The PCR conditions were as follows: initial activation at 95°C for 5 min, followed by 30 cycles of denaturation at 95°C for 40 sec, annealing at 55°C for 1 min, and extension at 72°C for 1 min, with a final extension cycle at 72°C for 5 min. PCR products were visualized after electrophoresis on a 2% agarose gel containing ethidium bromide at 135 V for 90 min. A 100 bp DNA ladder (Promega, USA) was used as a size marker. The classification of *E. coli* phylogenetic groups was performed by detecting the presence or absence of specific genes, namely *chuA*, *yjaA*, *uidA*, and *TspE4.C2*, using the primers listed in Table 1.

**Table 1.**
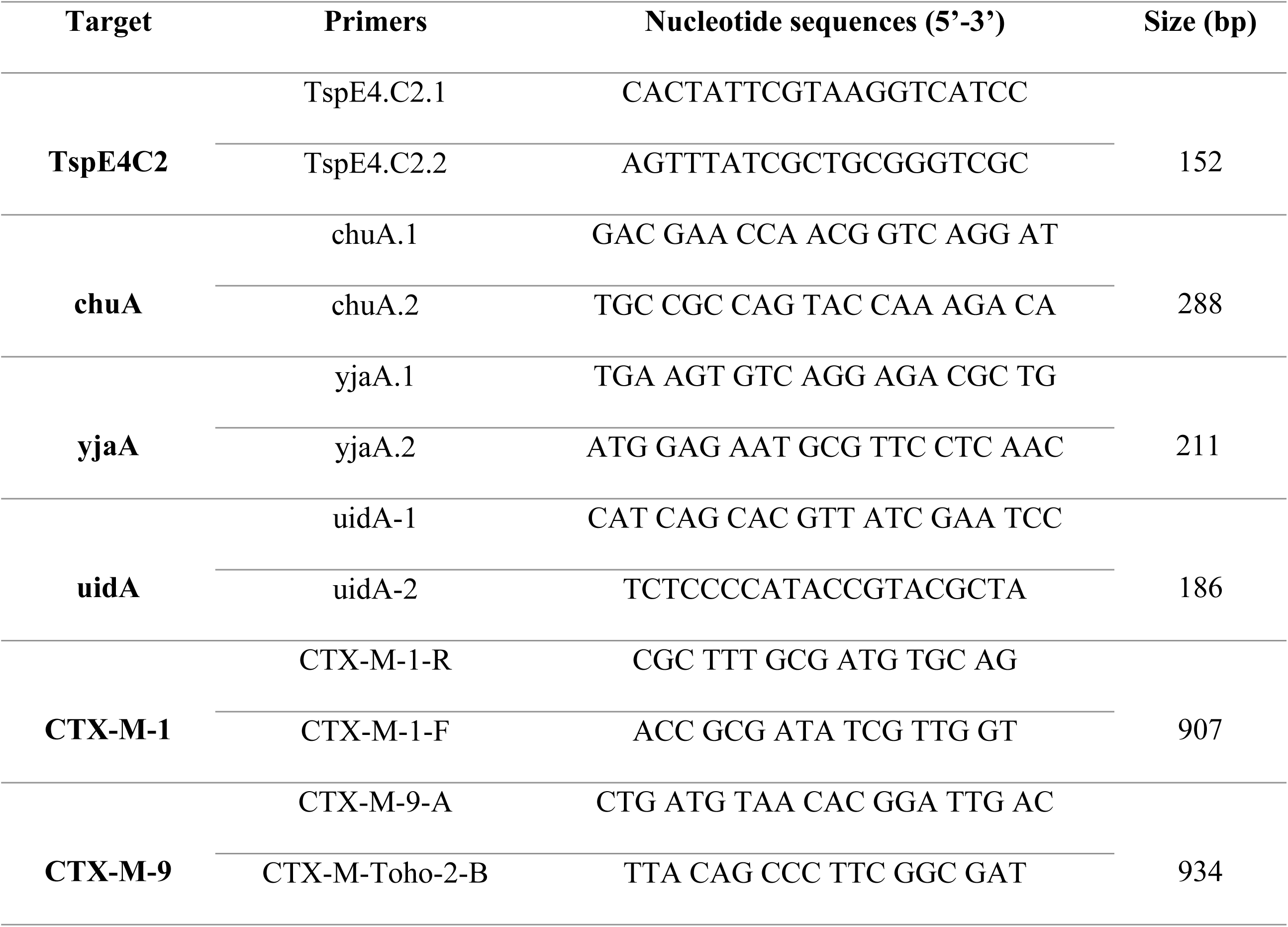
List of primers used for PCR-based phylogenetic grouping of *E. coli* isolates.

CTX-M genes (*CTX-M-1* group and *CTX-M-9* group) were detected by simplex PCR, adapting the method previously described by Batchelor et *al*. (2005) and De Champs et *al*. (2004), using the primers listed in Table 1. The PCR conditions were as follows: initial activation at 95°C for 5 min, followed by 30 cycles of denaturation at 95°C for 40 sec, annealing at 48°C for 40 sec (*CTX-M-1* group) and 52°C for 40 sec (*CTX-M-9* group), extension at 72°C for 1 min, and a final extension cycle at 72°C for 5 min. PCR products were visualized after electrophoresis on a 1% agarose gel containing ethidium bromide at 135 V for 45 min. A 100 bp DNA ladder (Promega, USA) was used as a size marker.

### Statistical analysis

Data were entered, recorded in the EPI info software version 7.2.2.6. Statistical analyses were performed using the Social Sciences statistical package (SPSS version 16.0). Word software was used for word processing. The comparison of percentages used the Mantel Haenszel Chi-square test or the Yates corrected Chi-square test in case of small numbers. The statistical significance level used was p = 0.05.

The sample size was determined using the formula of the binomial law described by Dagnelie (1998). By applying the No = p(1-p)×t2/m2 formula with a 95% confidence interval and a margin of error <<M>> of 5%, and p (58%) representing the prevalence of multidrug-resistant Enterobacteriaceae obtained by Ouedraogo et *al*. (2016), the estimated size of all samples combined was 1537 for this study.

## Results

### Demographic distribution of the study sample

Table 2 shows that 1,845 pathological samples were collected, including 599 (32.5%) from inpatients and 1,246 (67.5%) from outpatients. The number of positive culture samples for any microorganism was 32.25% [595/1,845; 95% CI: 30.16-34.42]. The mean age of patients was 33.46 ± 22.27 years [95% CI: 32.45-34.48]. The age group most affected was between 35 and 65 years. Male patients had a higher number of positive cases (58.70% [1083/1845; 95% CI: 56.44-60.93]) compared to female patients (41.30% [762/1845; 95% CI: 39.07-43.56]). Urine samples (cytobacteriological examination of urine) were the most common, comprising 52.19% of all cytobacteriological tests conducted [95% CI: 49.91-54.47].

**Table 2.**
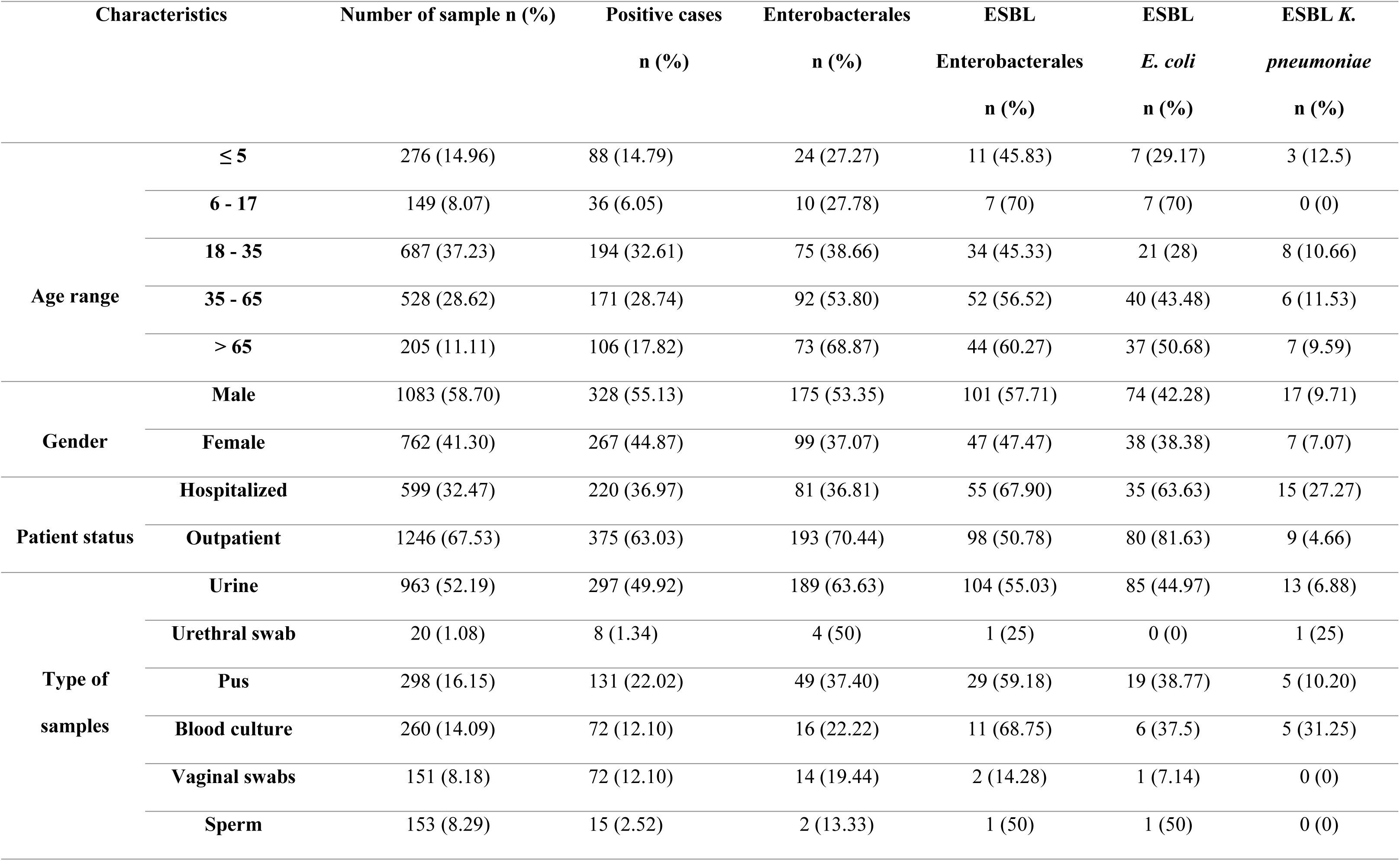
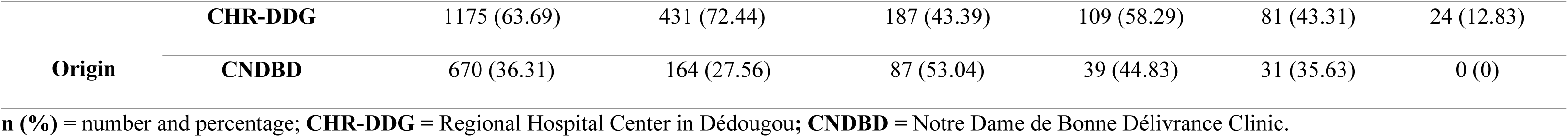
Demographic distribution of the study sample and prevalence of ESBL Enterobacterales.

### Isolation and identification of Enterobacterales

Table 3 illustrates that 274 *Enterobacterales* isolates were identified during the study. Of the isolated *Enterobacterales*, 70.80% [193/274; 95% CI: 64.65-75.78] were *E. coli* and 19.71% [54/274; 95% CI: 15.17-24.92] were *K. pneumoniae*. Other bacteria, such as *Citrobacter* sp. (1.46 %; 4/274), *Enterobacter cloacae* (2.19%; 6/274), *Proteus* sp. (3.65%; 10/274), and *Salmonella* sp. (2.19 %; 6/274), were also isolated. Urine samples contained the most *Enterobacterales*, with 78.31% [148/189; 95% CI: 15.17-24.92] of *E. coli* and 15.87% [30/189; 95% CI: 15.17-24.92] of *K. pneumoniae*.

**Table 3.** Distribution of isolated Enterobacterales species by sample type.

| Samples | Enterobacterales | Species |  |
| --- | --- | --- | --- |
|  |  | *E. coli* | *K. pneumoniae* |
| Urine n (%) | 189 (68.98) | 148 (78.31) | 30 (15.87) |
| Pus n (%) | 49 (17.88) | 25 (51.02) | 12 (24.5) |
| Blood n (%) | 16 (5.84) | 8 (50) | 6 (37.5) |
| Vaginal swabs n (%) | 14 (5.10) | 10 (71.42) | 4 (28.57) |
| Urethral swabs n (%) | 4 (1.46) | 2 (50) | 1 (25) |
| Sperm n (%) | 2 (0.07) | 1 (50) | 1 (50) |
| Total n (%) | 274 (100) | 194 (70.80) | 54 (19.71) |

N (%) = number and percentage

### Resistance profile of extended-spectrum β-lactamase-producing *E. coli* and *K. pneumoniae* isolates

Of the 274 *Enterobacterales* isolates confirmed, 162 (59.34%) [95% CI: 53.26-65.22] and 217 (79.20%) [95% CI: 73.90-83.85] were resistant to cefotaxime and amoxicillin + clavulanic acid, respectively, as shown in Table 4. Beta-lactam resistance phenotypes were detected in 148 (54.01%) [95% CI: 47.73-59.82] of the *Enterobacterales*, manifested by the appearance of the champagne cork, as shown in Figure 1. Among the species, the prevalence of ESBL-producing *E. coli* was 57.73 % (112/194), followed by *K. pneumoniae* (44.44 %; 24/54), *Citrobacter* sp (50 % 2/4), *E. cloacae* (83.33 % 5/6), *Proteus* sp. (40 %; 4/10), and *Salmonella* sp. (16.66 %; 1/6). A relatively low level of resistance to gentamicin (33.21%), fosfomycin (10.50%), nitrofurantoin (3.65%), imipenem, and ertapenem (1.46%) was observed in the isolates, as shown in Table 4. Similarly, low rates of resistance to fosfomycin (1.79%), nitrofurantoin (5.35%), and ertapenem (0%) were observed in ESBL *E. coli*, as shown in Table 4. Conversely, high resistance rates to nitrofurantoin (24.14%), gentamicin (70.83%), and fosfomycin (70.83%) were observed in ESBL *K. pneumoniae*, as shown in Table 4.

**Fig. 1.**
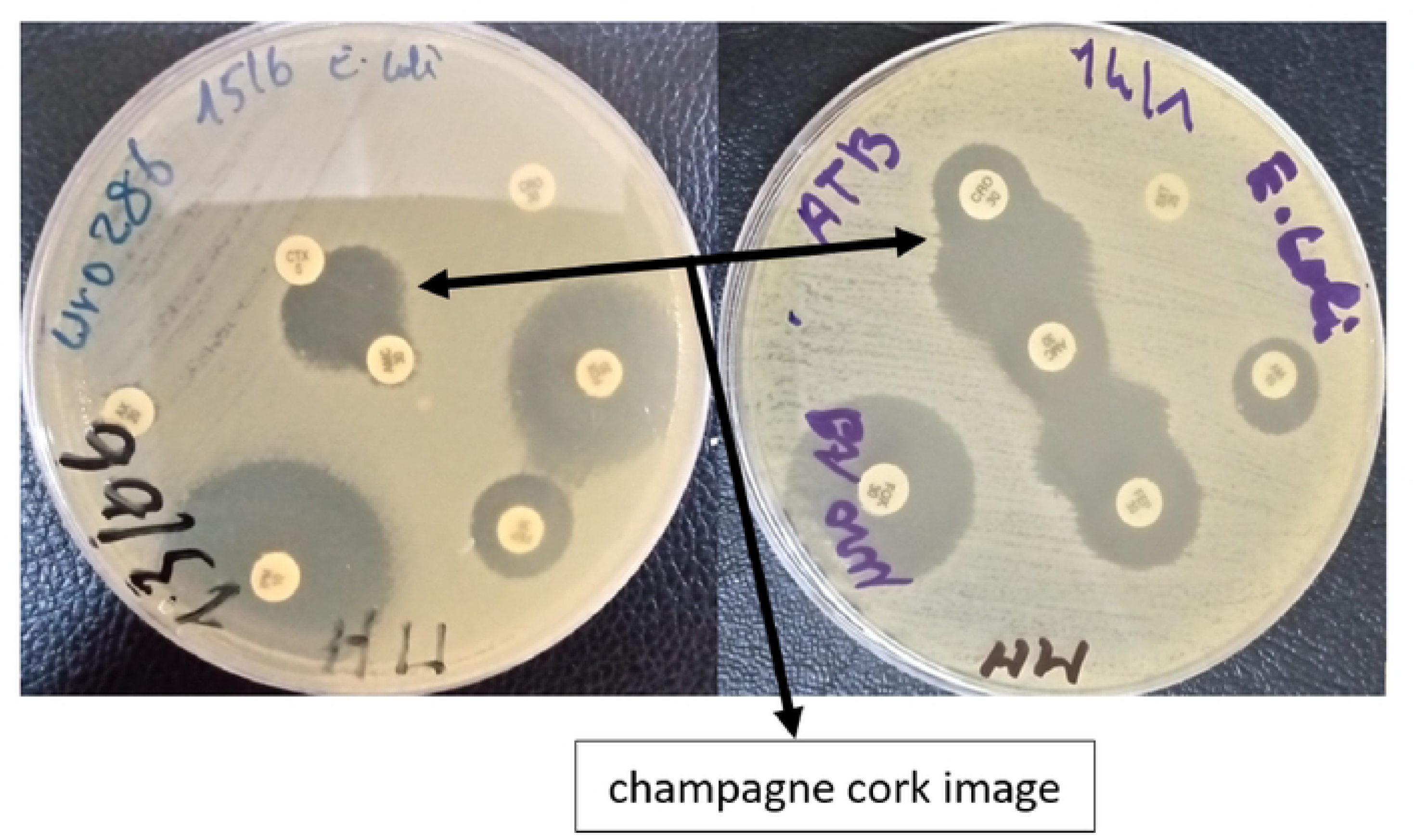
Phenotypic detection of ESBL production (Source: Coulibaly, 2024)

**Table 4.**
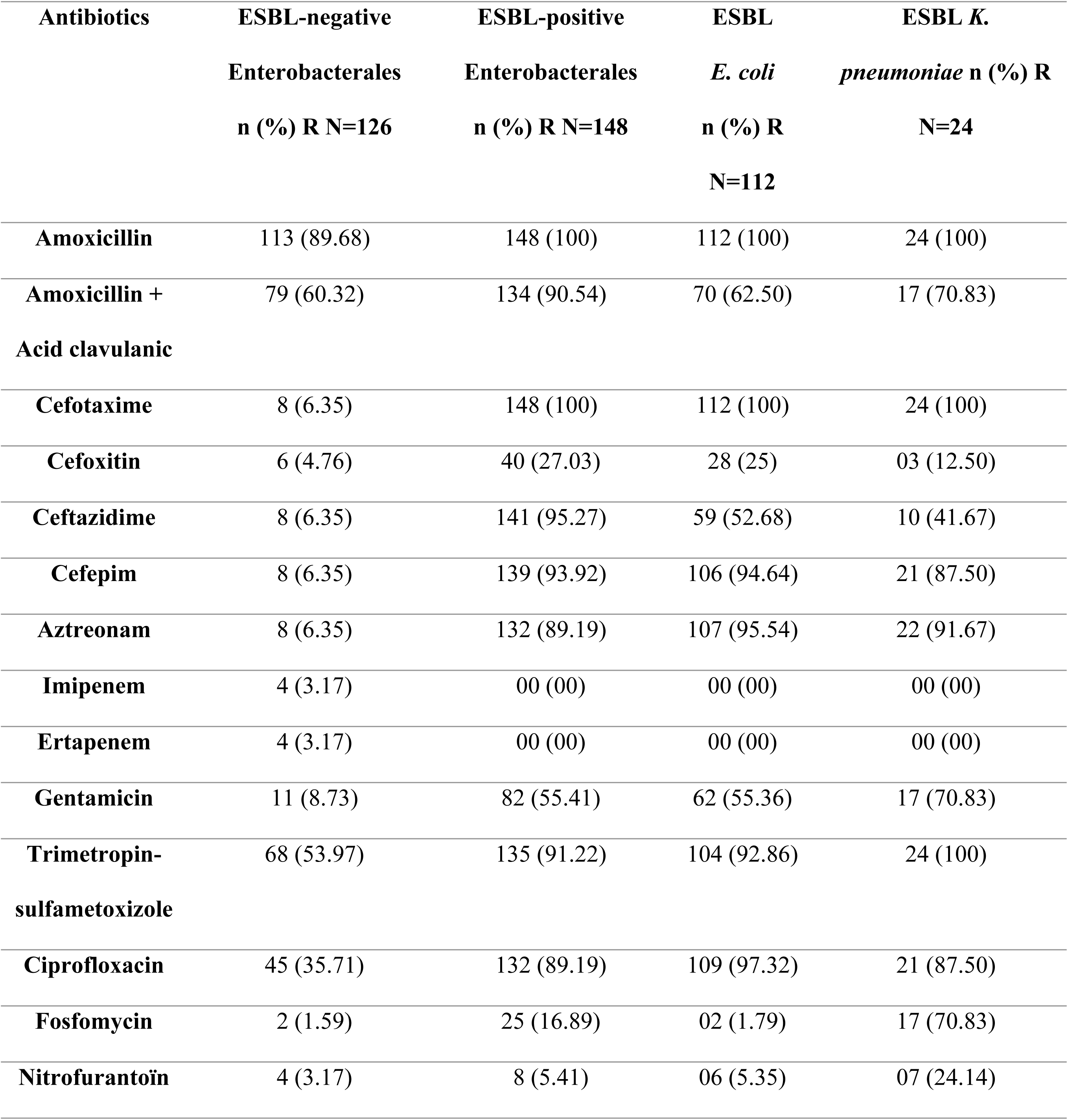
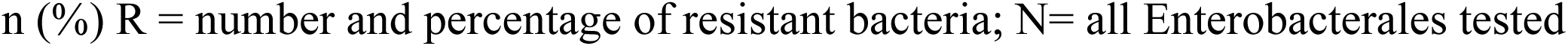
Antibiotic susceptibility of Enterobacterales.

| Antibiotics | ESBL-negative<br>Enterobacterales<br>n (%) R N=126 | ESBL-positive<br>Enterobacterales<br>n (%) R N=148 | ESBL<br>*E. coli*<br>n (%) R<br>N=112 | ESBL *K.*<br>*pneumoniae* n (%) R<br>N=24 |
| --- | --- | --- | --- | --- |
| **Amoxicillin** | 113 (89.68) | 148 (100) | 112 (100) | 24 (100) |
| **Amoxicillin +**<br>**Acid clavulanic** | 79 (60.32) | 134 (90.54) | 70 (62.50) | 17 (70.83) |
| **Cefotaxime** | 8 (6.35) | 148 (100) | 112 (100) | 24 (100) |
| **Cefoxitin** | 6 (4.76) | 40 (27.03) | 28 (25) | 03 (12.50) |
| **Ceftazidime** | 8 (6.35) | 141 (95.27) | 59 (52.68) | 10 (41.67) |
| **Cefepim** | 8 (6.35) | 139 (93.92) | 106 (94.64) | 21 (87.50) |
| **Aztreonam** | 8 (6.35) | 132 (89.19) | 107 (95.54) | 22 (91.67) |
| **Imipenem** | 4 (3.17) | 00 (00) | 00 (00) | 00 (00) |
| **Ertapenem** | 4 (3.17) | 00 (00) | 00 (00) | 00 (00) |
| **Gentamicin** | 11 (8.73) | 82 (55.41) | 62 (55.36) | 17 (70.83) |
| **Trimetropin-**<br>**sulfametoxizole** | 68 (53.97) | 135 (91.22) | 104 (92.86) | 24 (100) |
| **Ciprofloxacin** | 45 (35.71) | 132 (89.19) | 109 (97.32) | 21 (87.50) |
| **Fosfomycin** | 2 (1.59) | 25 (16.89) | 02 (1.79) | 17 (70.83) |
| **Nitrofurantoin** | 4 (3.17) | 8 (5.41) | 06 (5.35) | 07 (24.14) |

n (%) R = number and percentage of resistant bacteria; N= all Enterobacterales tested

### Detection of phylogenetic groups in *E. coli*

A total of 112 isolates of ESBL *E. coli* were grouped into four phylogroups, dominated by 47.32% [95% CI: 37.81-56.98] in group B2 and 42.86% [95% CI: 33.55-52.55] in group A, as shown in Figure 2 and Table 5.

**Fig. 2.**
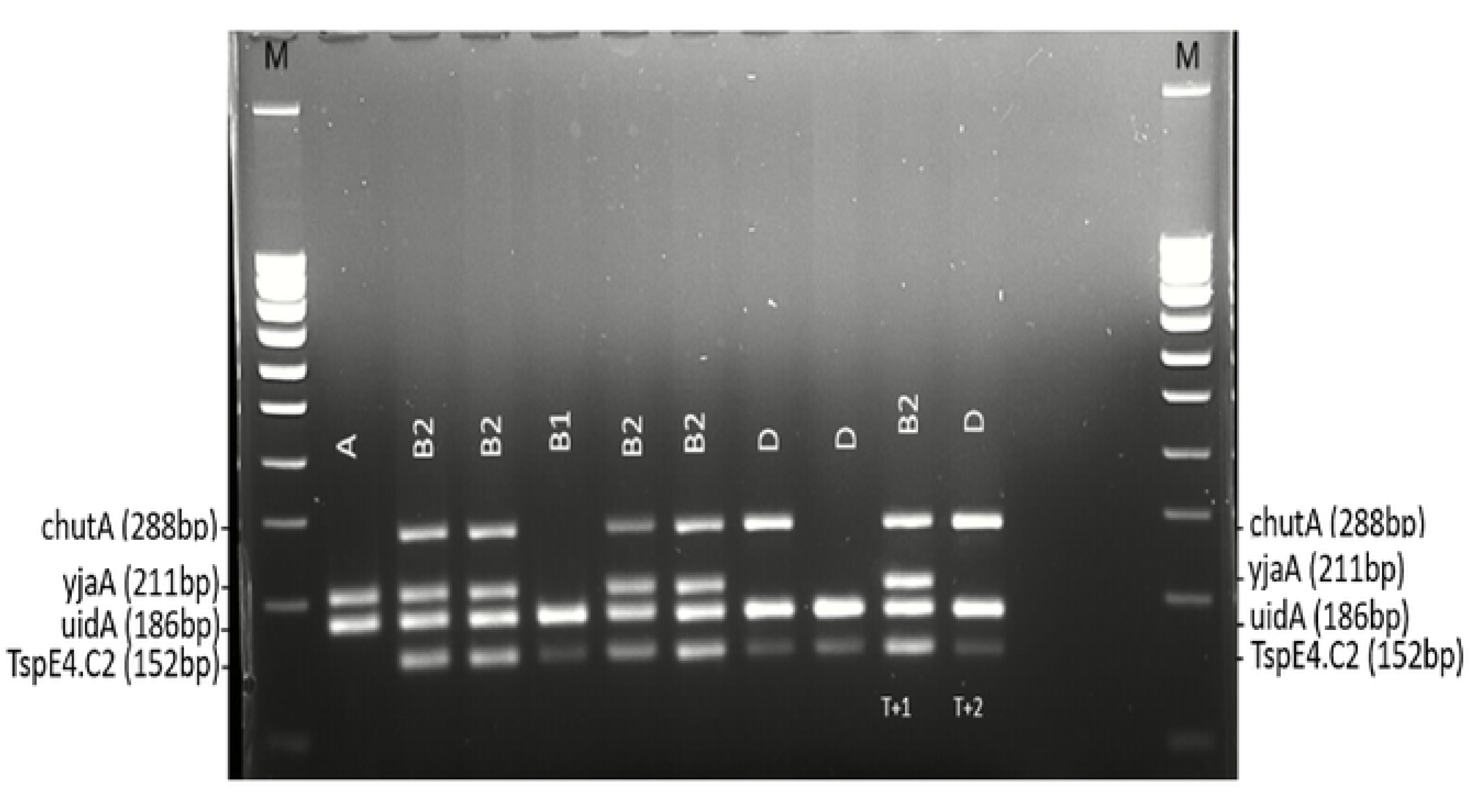
Quadruplex PCR for the detection of ESBL *E.coli* phylogroups. Legend: **M=** molecular weight (100 bp DNA ladder); T+I= positive control B2; T+2= positive control D; A-B1-B2-D= Phylogroups detected.

**Table 5.** Distribution of ESBL *E. coli* isolates according to the phylogroups detected.

| Phylogroups | Proportion N (%) | CI 95% Limits | *p*-value |
| --- | --- | --- | --- |
| A | 48 (42.86) | 33.55 – 52.55 | 0.156 |
| B2 | 53 (47.32) | 37.81 – 56.98 |  |
| B1 | 9 (8.04) | 3.74 – 14.71 | < 0.001 |
| D | 2 (1.79) | 0.22 – 6.30 |  |

### Identification of diverse CTX-M enzyme groups in *E. coli* and *K. pneumoniae*

Overall, Figures 3 and 4 show that the dominant CTX-M group detected among the ESBL-producing *Enterobacterales* was the CTX-M-1 group, with a frequency of 88.97% (121/136) [95% CI: 82.46-93.69; p < 0.05]. The CTX-M-9 group was detected in 11.03% (15/136) [95% CI: 6.31-17.54]. Among the ESBL-producing *E. coli*, 87.50% (98/112) [95% CI: 82.46-93.69] harbored the CTX-M-1 group, and 12.50% (14/112) [95% CI: 6.31-17.54] harbored the CTX-M-9 group. Similarly, in ESBL-producing *K. pneumoniae* isolates, the CTX-M-1 group and CTX-M-9 group were detected in 95.83% (23/24) [95% CI: 78.88-99.89] and 4.17% (1/24) [95% CI: 0.11-21.12], respectively (p < 0.05).

**Fig. 3.**
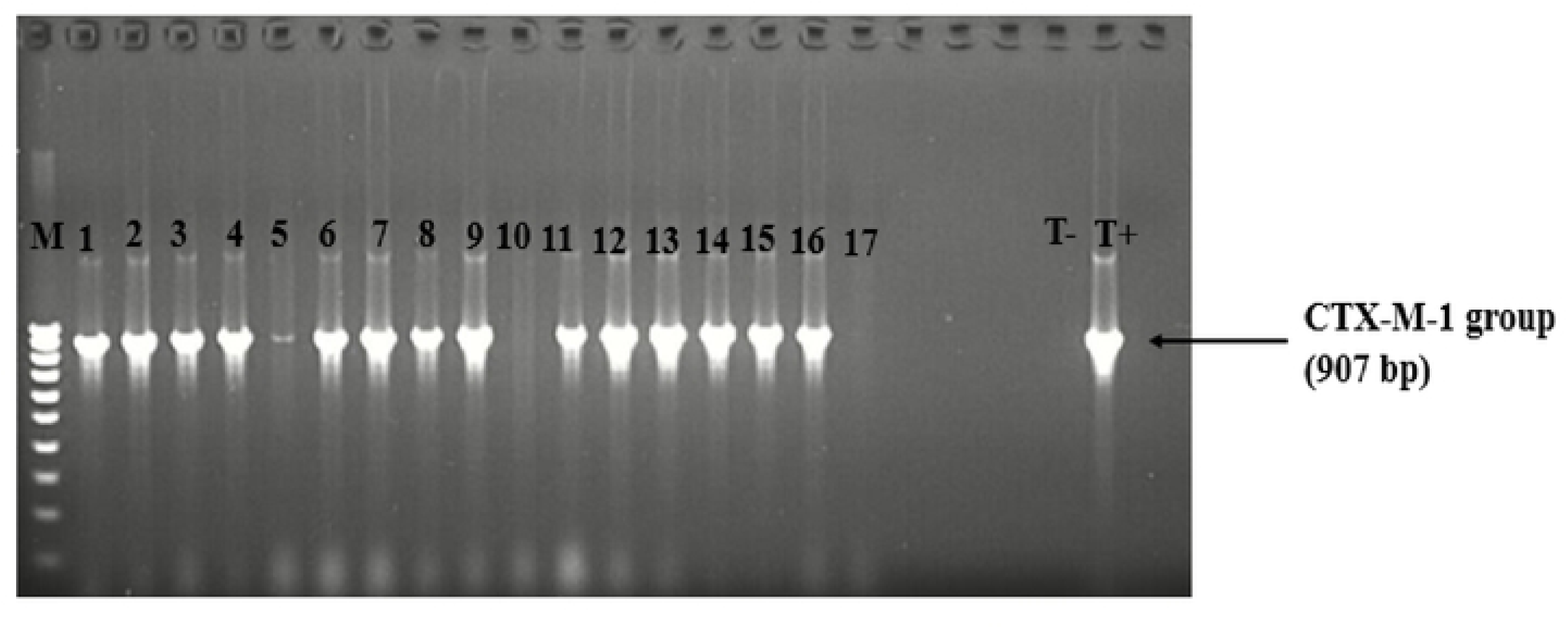
Gel image showing **the** detection of CTX-M-1 group in *E. coli* and *K. p11eumo11iae*. Legend: M = molecular weight (100 bp DNA Ladder); lane I to 5, 6 lo 9, 11 lo 16 = CTX-M-1 group positive; T+ = positive control; T-= negative control; 10 and 17 = CTX-M-1 group negative.

**Fig. 4.**
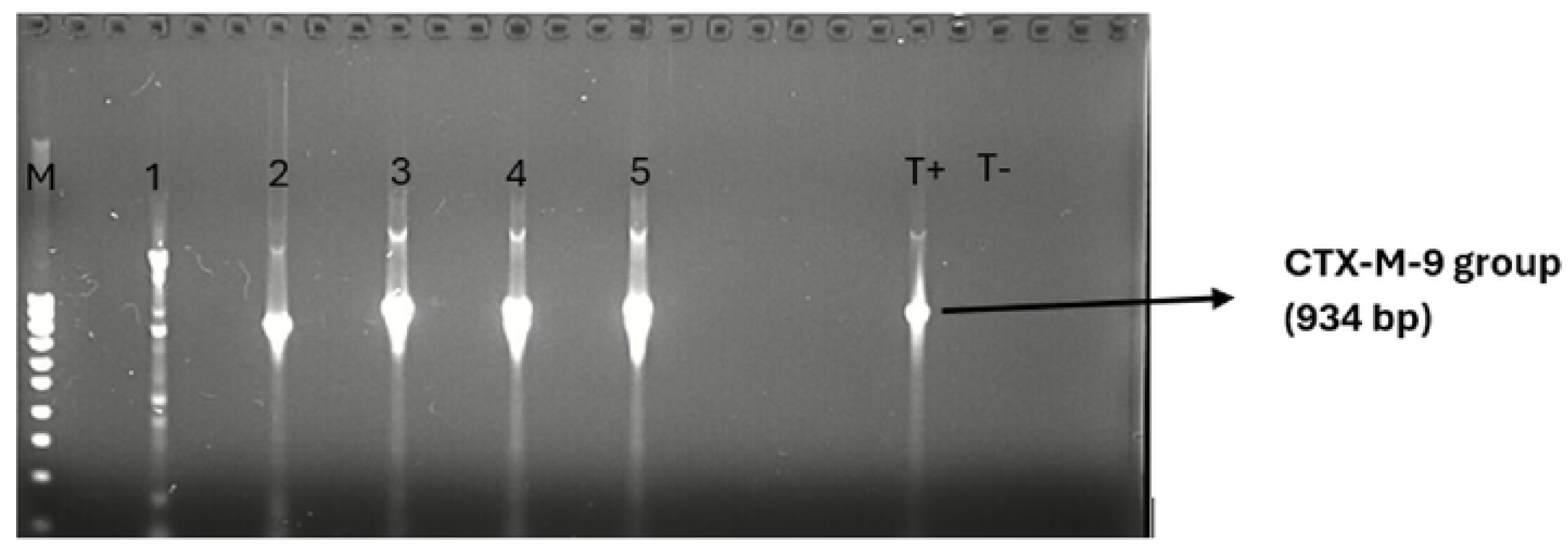
Gel image showing the detection of CTX-M-9 group in ESBL *E. coli* and *K. pneumoniae*. Legend: M =molecular weight (100 bp DNA Ladder); lane I to 5 = CTX-M-9 group positive; T+= positive control T-= negative control.

### Distribution of blaCTX-M genes within *E. coli* phylogroups

Figure 5 indicates that the distribution of resistance genes did not differ significantly between phylogroups (Phylogroup A: 91.67% of the CTX-M-1 group and 8.33% of the CTX-M-9 group; Phylogroup B2: 88.89% of the CTX-M-1 group and 11.11% of the CTX-M-9 group; p = 0.766). Moreover, within *E. coli* of the B1 phylogroup, the majority (88.89%; 8/9) harbored the CTX-M-1 group. As for *E. coli* of the D phylogroup, both harbored the CTX-M-1 group.

**Fig. 5.**
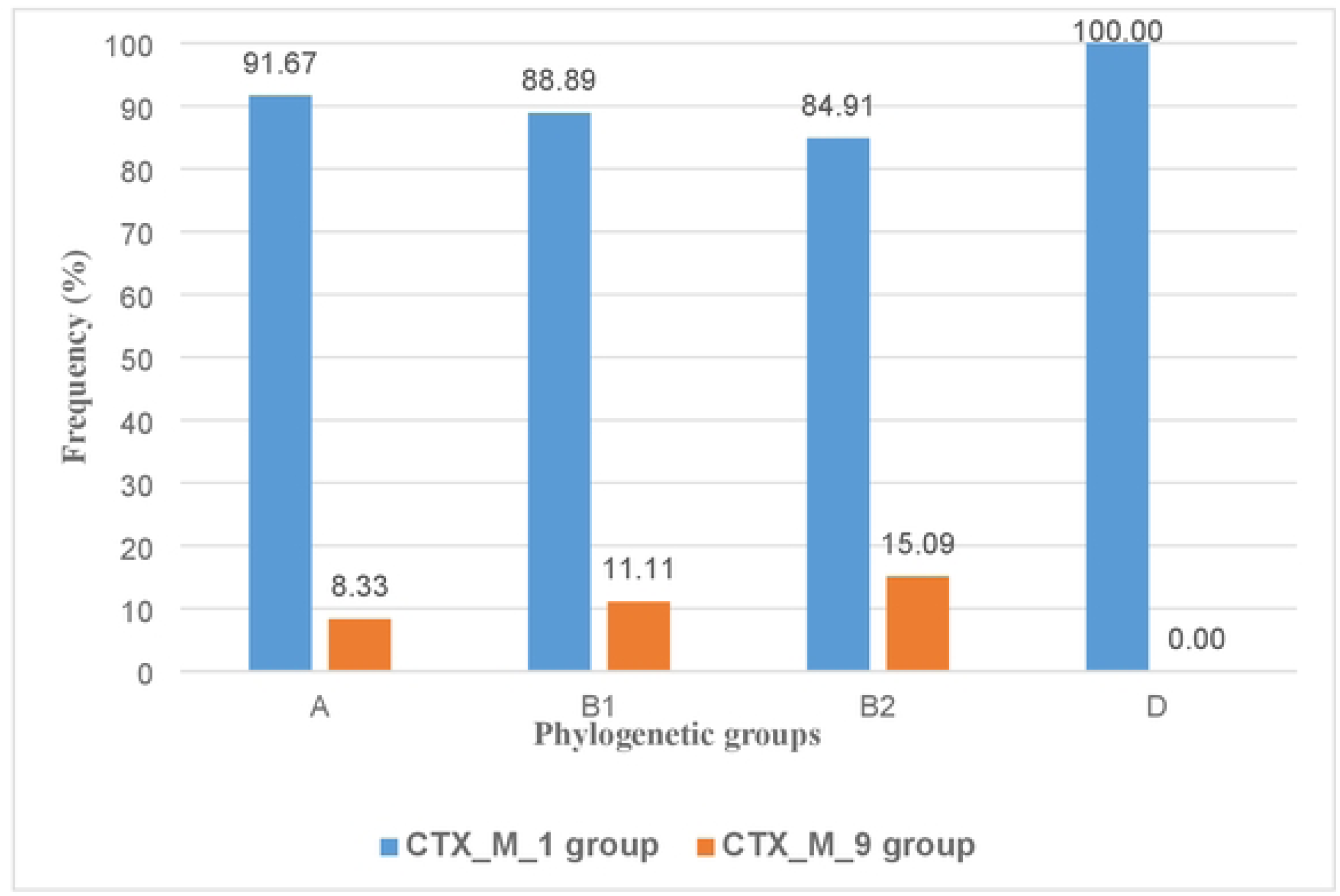
Distribution of *bla CTX-M* genes within ESBL-producing *E. coli* phylogroups.

## Discussion

Globally, there is a concerning surge in the emergence of ESBL-producing *Enterobacterales* [17]. In this study, 54.01% of *Enterobacterales* isolated from clinical samples were ESBL-producing isolates. When comparing studies from Burkina Faso, our results are lower than those of some studies [9,18], but higher than others [11,19–22], as well as internationally [3,8,23]. The disparity in prevalence could be attributed to factors such as sample type, sample size, or methodology applied. The high level of ESBL production by *Enterobacterales* in Burkina Faso is thought to be due to their constant and uncontrolled exposure to third-generation cephalosporins (cefotaxime, ceftriaxone, and cefixime), which are widely used in therapeutics. This widespread use leads to selection pressure and the development of multi-resistant isolates. This selective pressure is evident, as 59.34% of isolates in this study were resistant to cefotaxime. Taking into account the origin of the samples, 58.29% of ESBL *Enterobacterales* were identified at the RHC-DDG, compared with 44.83% at the CNDBD-DDG. This difference in prevalence could be explained by the fact that the CNDBD-DDG is used exclusively by outpatients, whereas the RHC-DDG serves as the region’s primary referral center for all types of patients. Additionally, a significant difference was observed in the prevalence of ESBL *Enterobacterales*, with 50.78% in outpatients compared to 67.90% in inpatients (p = 0.002), which aligns with the results of other studies [9]. Among outpatients, males were the most affected, comprising 68.75% [24], and the age groups most at risk were ‘35-65 years’ and ‘65 and over,’ with 36.63% and 38.61%, respectively (p = 0.0007). Regarding pathological samples, the production of ESBL by *Enterobacterales* was more prevalent in urine samples than in other sample types [9,20,25]. However, when considered individually, urine samples ranked third with a prevalence of 55.03%, following blood cultures and pus samples, which had respective prevalences of 68.75% and 59.18% [9]. Divergent results were obtained by Gnada et *al*. [11], who showed in their study that pus samples were the most infected by ESBL-producing isolates. Of the bacteria implicated in the production of the ESBL enzyme in our study, *E. coli* ranked first with 57.73 % of ESBL production, followed by *K. pneumoniae* with 44.44 %. This observation is supported by previous studies [9,11,17,25–27], which indicate that *E. coli* and *K. pneumoniae* are the main ESBL producers both locally and internationally. However, regarding the order of ESBL production, a study conducted in Nepal reported *K. pneumoniae* as the leading ESBL producer among other isolates [28]. It was also noted in this study that ESBL *E. coli* were much more frequently implicated in community infections than in hospital infections (69.56%) [95% CI: 60.29-77.80], and were correlated with age (CC: 0.164; p = 0.047) [9,29]. ESBL-producing *E. coli* and *K. pneumoniae* were found to be much more resistant to cefotaxime, cefepime, aztreonam, ciprofloxacin, and trimethoprim-sulfamethoxazole but sensitive to carbapenems (imipenem, ertapenem) as well as fosfomycin and nitrofurantoin, which are used in the treatment of urinary tract infections [9,27]. The level of sensitivity of ESBL isolates to carbapenems observed in our study contrasts with the findings of other authors [26,28], who reported that in their studies, *E. coli* and *K. pneumoniae* ESBL isolates showed a high rate of resistance to carbapenems. The high resistance of our ESBL isolates to the majority of ß-lactam antibiotics and other antibiotics (ciprofloxacin, trimethoprim-sulfamethoxazole) can be explained by the fact that in this region of Burkina Faso, urbanization is at a low level, and probabilistic prescriptions (sometimes by health workers or clandestine clinics) are more common, often without diagnostic advice from medical diagnostic laboratories. This brings us back to the issue of ‘self-medication,’ which is a leading cause of antibiotic resistance. Patients often seek care at health centers only with symptoms of advanced infections after prolonged exposure to self-medication. In addition, although the use of antibiotics in the animal production chain is often controlled by veterinary officers on prescription, some players (farmers and livestock breeders) frequently misuse and apply them without control in order to increase productivity and maintain animal health. Sometimes, the antibiotics used in veterinary medicine are equivalent to or belong to the same family as those used in human medicine. As a result, the consumption of food products of animal origin contaminated with residual antibiotics and sometimes with multidrug-resistant bacteria can pose serious public health risks [30]. On the other hand, the low resistance rates observed with carbapenems can be explained by the fact that these antibiotics are last resort, very expensive, and rarely prescribed. As for fosfomycin and nitrofurantoin, the preservation of their activity, likely due to their low usage, can be explained by their specific indications: they are reserved for urinary tract infections. Given this level of multi-drug resistance in our isolates at the phenotypic level, all 136 isolates of ESBL *E. coli* and *K. pneumoniae* harbored CTX-M genes, comprising 88.97% of the CTX-M-1 group and 11.03% of the CTX-M-9 group (p < 0.05). Our results are consistent with those of previous studies conducted in Burkina Faso [9,10] and internationally [3,4,8,22,24,28,31], which show that genes belonging to the blaCTX-M-1 group are most often responsible for the ESBL phenotype in clinical settings and are disseminated by *E. coli* isolates carrying blaCTX-M-15 (a variant of the blaCTX-M-1 group). However, other studies have shown that the bla-TEM gene is the most dominant gene compared to blaCTX-M [27,32,33]. The distribution of phylogenetic groups in ESBL-producing *E. coli* in this study was not statistically significant. Additionally, no significant correlation was observed between the phylogenetic groups of ESBL *E. coli* isolates and the dissemination of blaCTX-M-15 (p > 0.05). Furthermore, the two ESBL *E. coli* isolates, both isolated from urinary tract infections and assigned to phylogroup D (virulent extra-intestinal group), also carried the blaCTX-M-1 group. This is consistent with the findings of other authors [9]. Given the phylogenetic diversity observed within our ESBL *E. coli* isolates, it is unlikely that the dissemination of multidrug-resistant bacteria can be attributed to bacterial transmission between patients. Instead, it is more probable that the spread of plasmids carrying antibiotic resistance genes is responsible for this phenomenon, driven by selection pressure. Future studies will necessarily include investigations into multi-locus sequence typing, plasmid characterization, gene-encoded virulence factors, and our high-level cephalosporinase– and carbapenemase-producing *E. coli* isolates. The main limitation of this study is the small number of health facilities, which makes generalization difficult. Further studies involving a larger number of health facilities than those considered in this study would be necessary in the Boucle du Mouhoun region once the safety situation has improved. These studies could include the whole genome sequencing of bacterial isolates.

## Conclusion

In conclusion, the results of our findings revealed a worrying frequency of ESBL-producing *E. coli* and *K. pneumoniae* in this region of Burkina Faso. Molecular analysis revealed significant circulation of the genes CTX-M-1 group. These isolates were much more resistant to ß-lactam antibiotics (except carbapenems), quinolones, often aminoglycosides and other antibiotics commonly used by people in the region. Measures (implementation of antimicrobial stewardship and infection prevention and control) must be adopted to control antimicrobial resistance to preserve antibiotics that are still effective and improve the quality of care in health centres at the local and national levels.

## Data Availability

All relevant data are within the manuscript and its Supporting Information files.

S/O

## Acknowledgements

We thank the hospital authorities for their agreement and the staff of the laboratory department of both the RHC-Dédougou and CNR-Clermont Ferrand for their support respective in obtaining the samples and molecular characterization of bacterial isolates.

## Author contributions

H.C. is responsible for initiating the study and analysing the data, preparation of all figures and tables, and writing of the manuscript. Laboratory examinations were conducted by H.C. under the supervision of D.S.K., O.T., A.B.K., A.H.M., H.K., D.O., D.B., R.F., N.B., R.B. and R.D. Data analysis and manuscript preparation were overseen by R.B. and R.D. All the authors contributed to and reviewed the final manuscript.

## Consent for publication

All authors have read and approved the final version of the manuscript for publication.

## Availability of data and materials

All data obtained are available within the article.

